# School and meal characteristics associated with plate waste in K-12 cafeterias in the United States

**DOI:** 10.1101/2024.02.06.24302396

**Authors:** Eunice S. Adjapong, Kathryn E. Bender, Sophia Schaefer, Brian E. Roe

## Abstract

Food wasted in primary and secondary education institutions creates nutritional losses, financial inefficiencies, and environmental degradation. While there is some evidence of how particular interventions within schools may influence the amount of waste created, there is little recent information about typical levels of food waste observed in U.S. schools and how these correlate with school and meal characteristics. We analyze data reported by more than 100 schools from 24 states as part of the World Wildlife Fund’s Food Waste Warriors project and identify how plate and beverage waste from school lunches are associated with school and meal service characteristics. We find schools that permit students to choose their own amount of milk report 76% less milk waste than those reliant upon individual milk cartons while schools that implement at least one non-curricular intervention (e.g., a table where students can share unopened) report significantly less produce waste than other schools. We confirm several patterns observed or hypothesized in the literature, including more waste generated by younger students and during the earliest and shortest lunch periods. We document several novel associations including more plate waste at smaller schools, during winter months and in the Northeast region. We find several nuanced patterns of waste related to the prevalence of free and reduced meal service and whether all meal elements are offered versus served. While this study cannot support rigorous evaluation of intervention effectiveness, it provides insights into school and program characteristics that may pose challenges for schools interested in reducing student plate waste.

## Introduction

Federal and local governments invest billions of dollars per year in school meal programs to ensure that school-age children in the United States receive the nutrition needed to perform well in and outside of school (USDA 2023). When children leave food and beverages unconsumed during meals (plate waste), financial and natural resources are wasted, which undermines the goals of school meal programs while contributing to environmental degradation associated with wasted food. After reviewing studies of plate waste in primary and secondary (K-12) school cafeterias from the 1990’s, Buzby and Guthrie (2002) estimated that as much as 12 percent of the calories served as part of these programs were wasted, which was valued around $600 million, while studies using US data from the 2014/15 academic year estimate that 21% of served calories became plate waste (USDA 2019).

Buzby and Guthrie (2002) identified several possible reasons for cafeteria plate waste including that some meals are scheduled when children are not hungry (e.g., very early lunch periods or lunch periods before rather than after recess), deviations between child food preferences and meal content, and the inability to customize portion sizes to children’s appetites, among others. Cohen et al.’s (2021) review of more recent literature on school meal consumption points to several additional reasons for excessive plate waste including short lunch periods and limited options for individual meal components.

Despite significant changes in K-12 school meal programs over time, there has been limited analysis of plate waste patterns across U.S. schools which hampers identification of current inefficiencies and potential responses to reduce plate waste in school meal programs. Other than the large-scale USDA-administered studies summarized in Buzby and Guthrie (2002), Potamites and Gordon (2010) and USDA (2019), most research on U.S. school cafeterias has gathered data from a small number of schools in a limited geographic area, often with the intent to evaluate a specific intervention or policy change (e.g., Byker et al. 2014, Byker Shanks et al. 2021, Byker Shanks et al. 2023, Cohen et al. 2013, Blondin et al. 2014, Hakim and Meissen 2014, Wilkie et al. 2015, Sharma et al. 2021, Burg et al. 2021, Hass et al. 2014, Capps et al. 2019; see Cohen et al. 2021 for a systematic review). However, to our knowledge, there have been no studies analyzing waste data from schools drawn from throughout the United States since the USDA (2019) covering the 2014/15 school year.

Our work fills this gap by utilizing data gathered from World Wildlife Fund’s (WWF) Food Waste Warriors program, which encourages schools across the United States to conduct audits of school cafeteria plate waste to teach students about food waste and its environmental impacts on the food system and upload the results to a centralized database. Between 2018 and 2023, staff and volunteers from 134 schools in 24 states took advantage of this opportunity and provided data in a standardized format about the levels of plate waste they recorded, school features, and characteristics of their cafeteria and meals service. We analyze nearly 500 observations of the average plate waste per student, both in total and for four constituent categories (milk, entrees and sides, produce, and other liquids) and use regression analysis to identify associations between waste levels and the characteristics of the school and meals served.

## Materials and Methods

Data were collected by staff and volunteers at individual schools throughout the United States who participated in the World Wildlife Fund’s Food Waste Warriors program and entered results from a standardized audit process (USDA, EPA and University of Arkansas, undated) into the Food Waste Warriors Dashboard (WWF 2023). The audit prescribes methods for consistently collecting and measuring plate waste, including the use of an electronic kitchen scale or luggage scale to assess waste quantities. Some schools entered results from a single audit while other schools entered results from multiple audits which could consist of audits on several different days, across varying lunch periods on the same day, across varying subgroups of students or combinations therein. Each observation is derived from the audit of plate and non-water beverage waste at a single school and then expressed on a per student served basis. Waste is reported in total and separately for produce (fruits and vegetables), all other (non-produce) wasted food items (e.g., entrees and non-produce sides), milk, and, when applicable, all other non-water liquids (e.g., juice and soup). Data from an average of 3.5 days observed in 134 schools from 24 different states across all four regions were collected between August 2018 and May 2023 and were available for analysis. Eleven observations were excluded because the reported food waste figures represent data from 10 or fewer students. The school-level free or reduced lunch enrollment percentages were collected from state websites that published such data.

Sample summary statistics are listed in Table 1. While the sample consists of schools who took the initiative to audit and upload data (a self-selected convenience sample), Table 1 reveals a broad array of institutions who participated, including 45% with a majority of students receiving free or reduced-price meals and 71% who have not implemented any interventions to reduce school food waste. The sample skews towards elementary schools (just over half) and towards schools in western states (44%) with about twice as many observations occurring prior to the start of the COVID pandemic. The modal and median school provides a single audit, though several schools provided many audits (up to 27 in one case) and, hence, the average number of audits per school is nearly 4.

**Table 1.** Summary Statistics.

| VARIABLES | Mean or % | Std. Dev. |
| --- | --- | --- |
| Lunch Start Time |  |  |
| < 11 am | 26.5% |  |
| 11:00 – 11:59 | 27.8 |  |
| Noon or later | 27.6 |  |
| Not reported | 18.2 |  |
| Length of Lunch |  |  |
| < 30 mins | 9.0 |  |
| 30 mins | 28.8 |  |
| > 30 mins | 5.7 |  |
| Not reported | 56.5 |  |
| Offer vs. Serve |  |  |
| Serve | 23.1 |  |
| Offer | 65.3 |  |
| Not reported | 11.6 |  |
| Free & Reduced Lunch |  |  |
| <25% | 20.8 |  |
| 25-49% | 20.8 |  |
| 50-74% | 17.6 |  |
| 75% or more | 27.1 |  |
| Not reported | 13.7 |  |
| Grades Served |  |  |
| Elementary | 54.7 |  |
| Middle | 33.5 |  |
| High | 9.2 |  |
| Not reported | 2.7 |  |
| School Size |  |  |
| <500 students | 88.0 |  |
| 500 – 999 students | 10.6 |  |
| 1000 or more students | 1.4 |  |
| Milk Service Method |  |  |
| In cartons |  |  |
| By dispenser or jug |  |  |
| Not reported |  |  |
| School in WWF Program (yes=1) | 55.9 |  |
| Interventions Implemented |  |  |
| None | 70.6 |  |
| 1 intervention (curricular) | 9.8 |  |
| > 1 intervention (curricular + other) | 10.4 |  |
| 1 intervention (non-curricular) | 6.5 |  |
| > 1 intervention (non-curricular) | 2.7 |  |
| Time Period |  |  |
| Pre-COVID | 63.3 |  |
| During COVID (Autum '20 – Spring '21) | 2.0 |  |
| Post COVID (Summer '21 or later) | 34.7 |  |
| Season |  |  |
| Spring | 37.6 |  |
| Summer | 1.2 |  |
| Autum | 15.9 |  |
| Winter | 45.3 |  |
| Region |  |  |
| Northeast (CT, MD, NH, NY, VT) | 9.2 |  |
| West (AK, AZ, CA, CO, HI, OR, WA) | 44.1 |  |
| Midwest (IL, IN, NE, OH, WI) | 29.0 |  |
| South (FL, GA, KY, LA, NC, TN, VA) | 17.8 |  |
| Number of entrees served | 2.457 | 1.596 |
| Number of previous audits | 3.855 | 5.133 |
| Total Plate + Beverage Waste | 0.290 | 0.181 |
| Produce waste | 0.101 | 0.088 |
| Non-produce food waste | 0.109 | 0.125 |
| Milk waste | 0.088 | 0.065 |
| Other liquid waste | 0.013 | 0.022 |

Notes: N=490 except for produce waste (N=450), non-produce waste (N=482), milk waste (N=456), and other liquid waste (N=328).

The distribution of the total waste is depicted in Figure 1 with separate box plots for elementary and all other schools (middle schools, high schools and schools who did not report grade level). To understand the school and meal level characteristics that are correlated with the observed waste levels, we regress these characteristics on the natural logarithm of total waste and each of the four waste subcategories using Stata (version 18.0). The distribution (Fig. 1) is clearly skewed with a long right tail (multiple outliers exceeding the upper whisker), which motivates our natural logarithm transformation of the waste levels for the purposes of regression analysis. Statistical significance is set at the 5% level with results featuring *p*-values between the 0.05 and 0.10 deemed marginally significant. We base all statistical tests on robust standard errors for the regression estimates.

**Figure 1.**
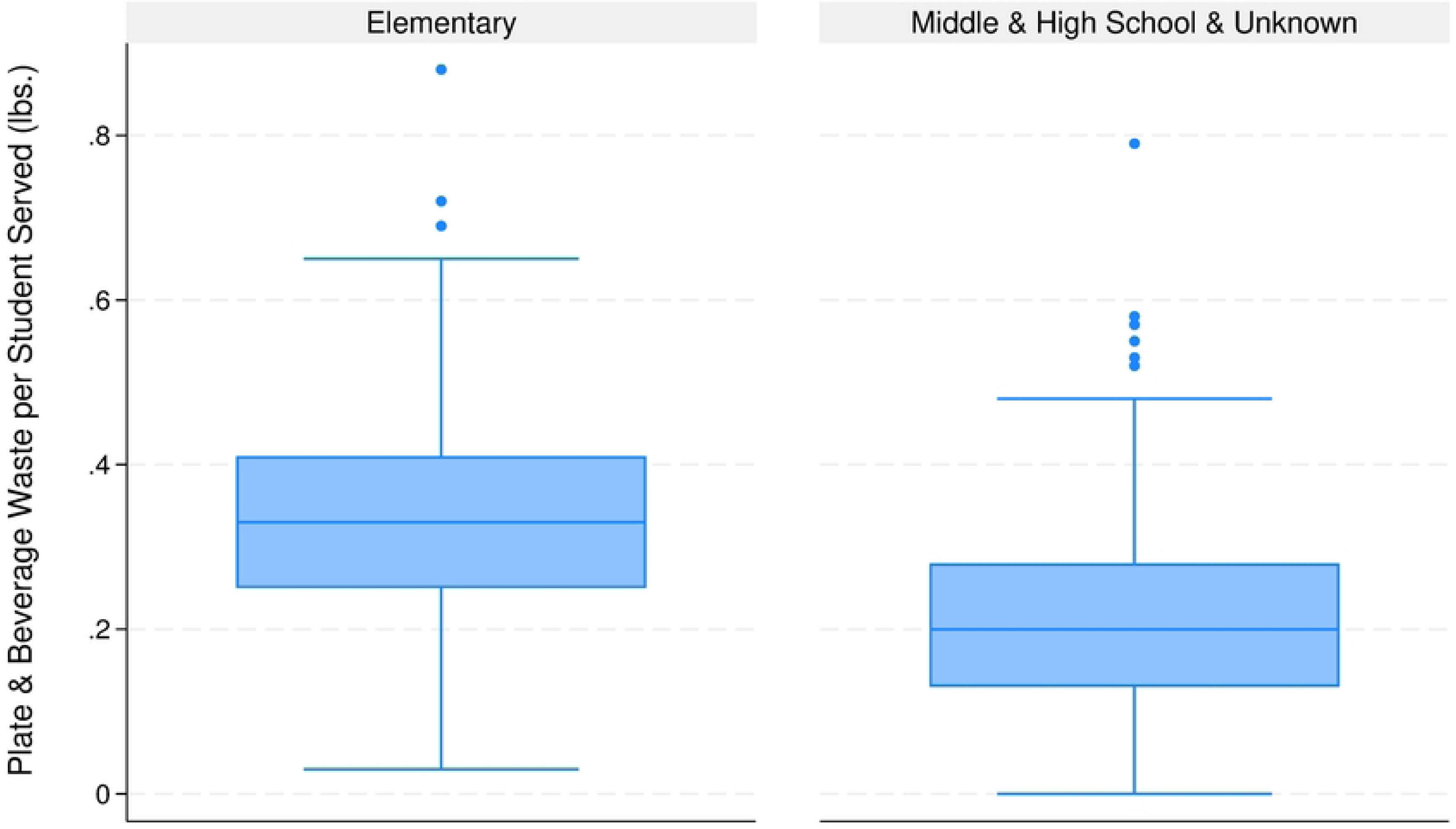
Box Plot of Plate and Beverage Waste per Student for Elementary and Other Schools. Scale truncated at 1 lb. per student, which omits 1 elementary observation (1.08 lbs./student) and 2 other school observations (1.35 and 2.39 lbs./student).

## Results

Average waste among the sample schools is 0.29 pounds per student per meal, which includes roughly equal amounts of produce waste (0.10), non-produce (e.g., entrée and non-produce sides) waste (0.11 lbs./student), and milk waste (0.09) with a small amount of other liquid waste (0.01). As confirmed in Table 2, plate waste is significantly greater in elementary schools with 52% more total waste than other schools. Most schools provide waste amounts by meal component, permitting assessment of association among waste streams (e.g., Figure 2 depicts a positive association between produce and non-produce waste within a school) with positive correlations between produce and non-produce (*r* = 0.079, *p*=0.097), produce and milk (*r =* 0.201, *p*<0.001) and milk and non-produce waste (*r* = 0.097, *p*=0.039). We can also confirm that significantly more waste is created in elementary schools for each of the three major meal components (Table 2).

**Figure 2.**
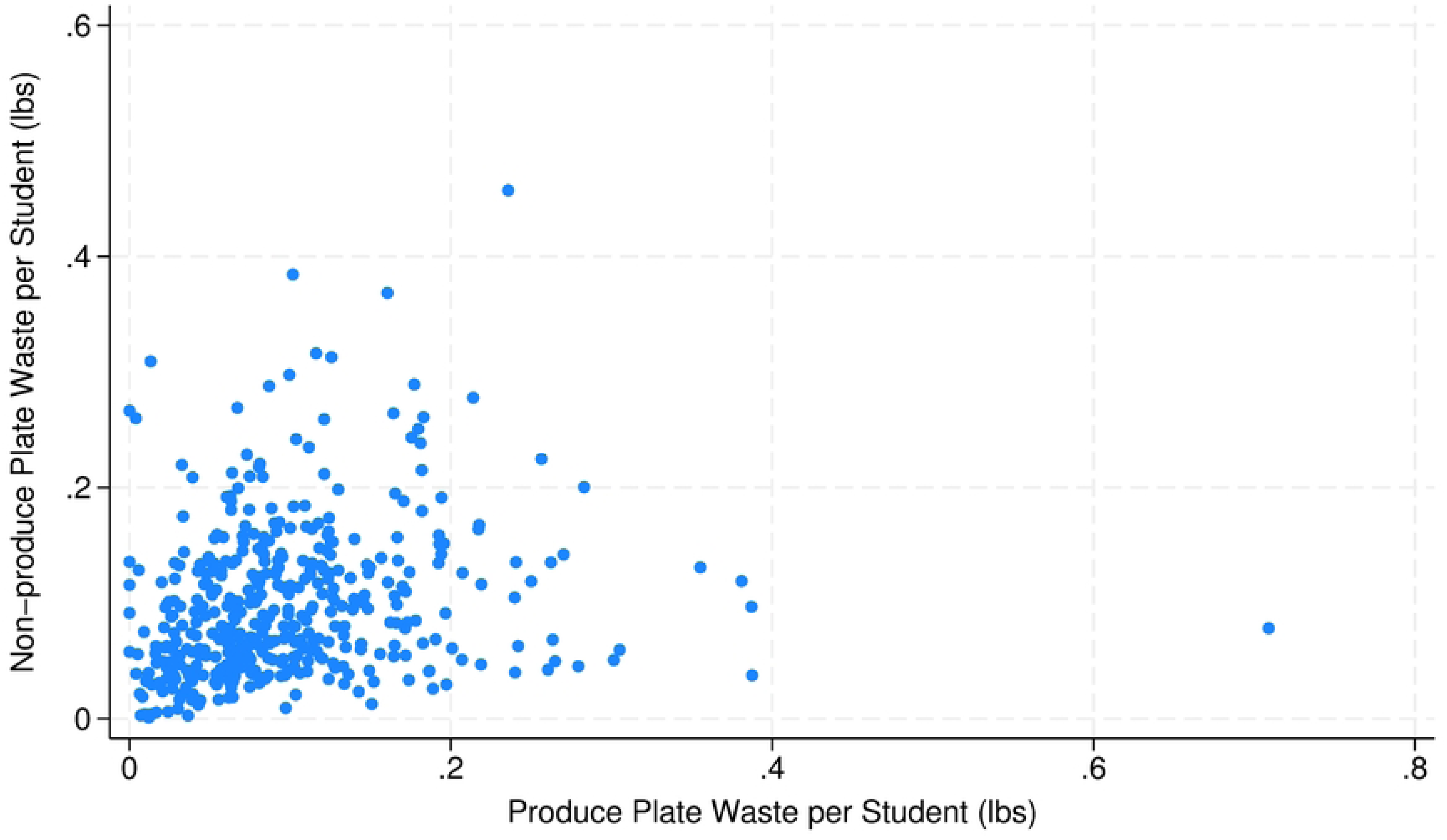
Scatter Plot of Plate Waste from Produce and Non-produce Foods (N=448). Axis truncated for purposes of visualization, omitting two observations with larger values.

**Table 2.** Average Plate and Beverage Waste by Age Group (lbs./student/meal).

| Variables | Elementary | All Other |  |
| --- | --- | --- | --- |
| Total Plate + Beverage Waste | 0.343<br>(0.008) | 0.226<br>(0.014) | n=490<br>$p < 0.001$ |
| Produce waste | 0.113<br>(0.004) | 0.085<br>(0.007) | n=450<br>$p < 0.001$ |
| Non-produce food waste | 0.123<br>(0.005) | 0.092<br>(0.011) | n=480<br>$p = 0.008$ |
| Milk waste | 0.111<br>(0.0040) | 0.058<br>(0.003) | n=456<br>$p < 0.001$ |

*Notes:* Standard errors in parentheses. $p$ -values are for the $t$ -test statistics of the null hypothesis of equality between the column figures. All Other schools include middle schools, high schools and schools that did not report grade level. No significant differences were identified between middle and high school averages other than for milk: middle school's reported 0.064 vs. 0.032 lbs./student for high schools ( $p < 0.001$ ).

The regression results for total food and beverage waste are presented in Table 3 for all schools and for two groups of schools (elementary and all other schools) while the regression results for three key subcomponents (non-produce, produce and milk with other liquid waste omitted due to fewer observations and less variation) pooled across all schools are presented in Table 4. The models generally display good fit with R^2^ statistics ranging from 0.585 (total waste from middle and high schools) to 0.376 (produce waste). We focus on presenting results on total waste and highlight when patterns differ by grade level or by subcomponent.

**Table 3.**
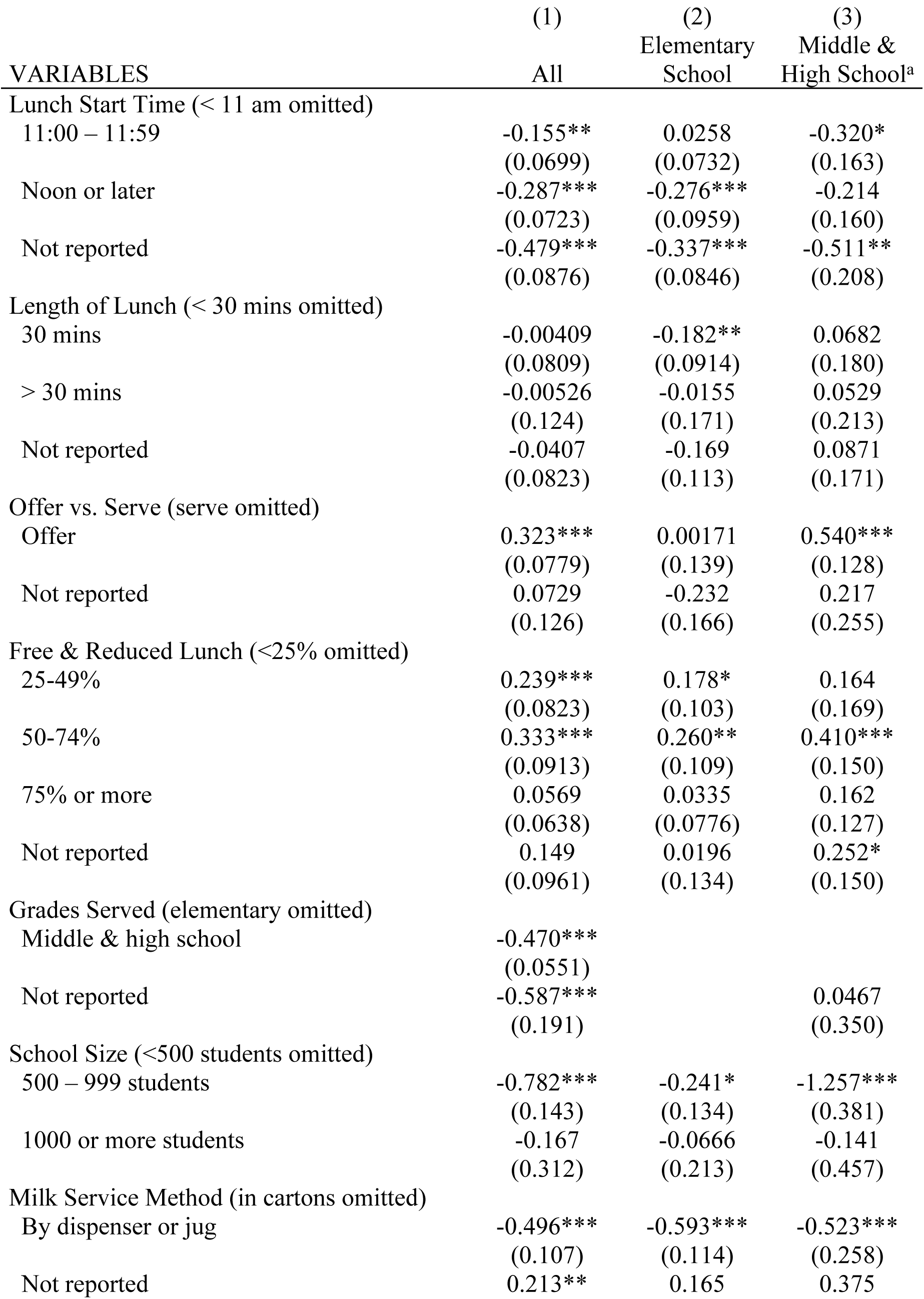
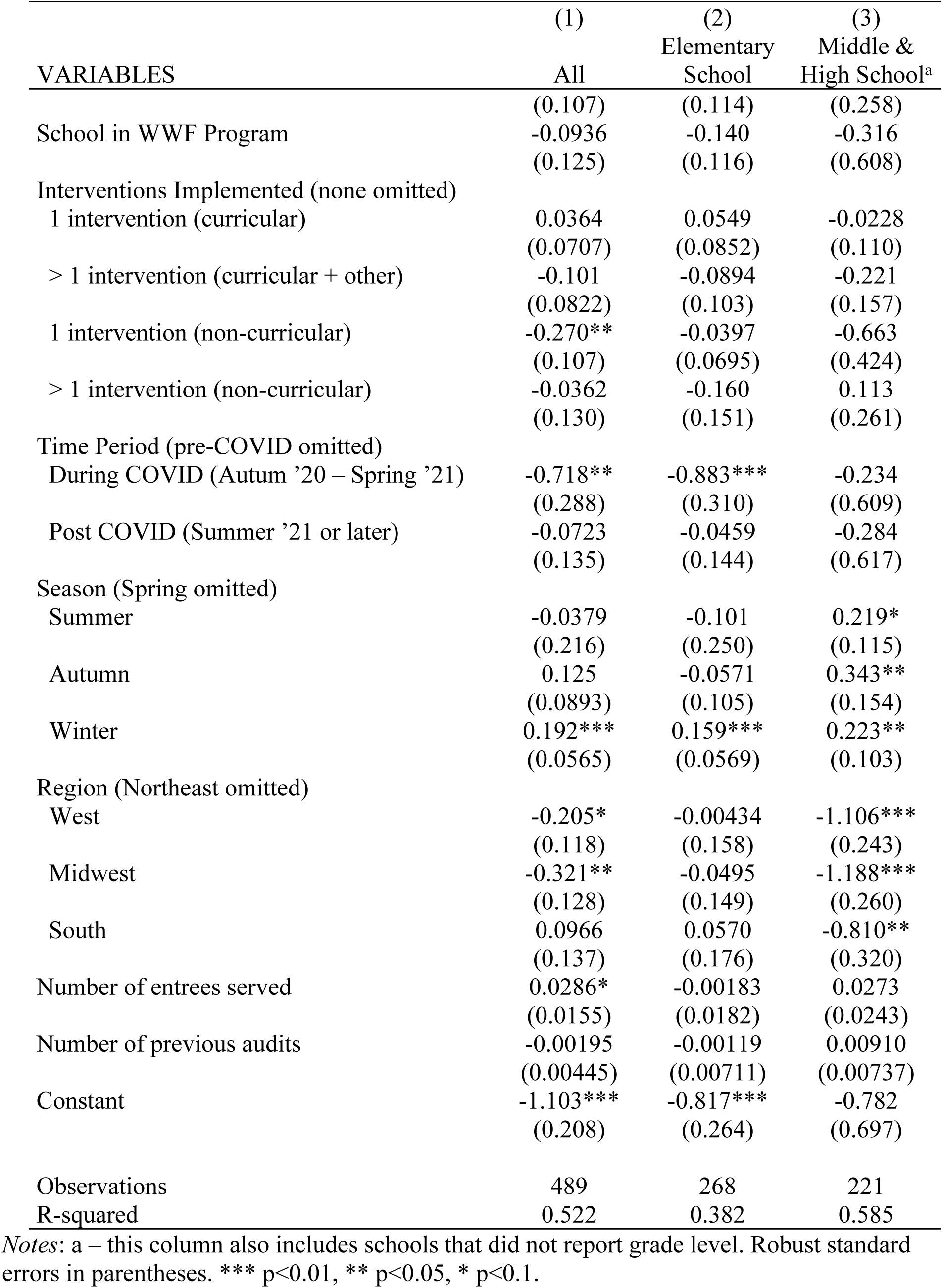
Total Plate and Beverage Waste Regression Results.

**Table 4.** Waste by Food Type Regression Results.

| VARIABLES | (1)<br>Non-produce | (2)<br>Produce | (3)<br>Milk |
| --- | --- | --- | --- |
| Lunch Start Time (< 11 am omitted) |  |  |  |
| 11:00 – 11:59 | -0.294***<br>(0.106) | -0.179<br>(0.128) | -0.172*<br>(0.0925) |
| Noon or later | -0.521***<br>(0.117) | -0.132<br>(0.134) | -0.270***<br>(0.0933) |
| Not reported | -0.506***<br>(0.129) | -0.509***<br>(0.149) | -0.383***<br>(0.125) |
| Length of Lunch (< 30 mins omitted) |  |  |  |
| 30 mins | 0.107<br>(0.120) | 0.133<br>(0.124) | -0.0495<br>(0.107) |
| > 30 mins | 0.105<br>(0.189) | -0.194<br>(0.190) | 0.103<br>(0.183) |
| Not reported | -0.0511<br>(0.130) | 0.0592<br>(0.116) | -0.310***<br>(0.116) |
| Offer vs. Serve (serve omitted) |  |  |  |
| Offer | 0.512***<br>(0.108) | 0.425***<br>(0.109) | 0.0540<br>(0.115) |
| Not reported | 0.205<br>(0.170) | 0.0514<br>(0.191) | -0.0800<br>(0.190) |
| Free & Reduced Lunch (<25% omitted) |  |  |  |
| 25-49% | 0.0960<br>(0.126) | 0.415***<br>(0.120) | 0.109<br>(0.113) |
| 50-74% | 0.0261<br>(0.120) | 0.629***<br>(0.134) | 0.147<br>(0.127) |
| 75% or more | -0.234**<br>(0.0978) | 0.241**<br>(0.0974) | 0.228**<br>(0.0974) |
| Not reported | 0.177<br>(0.133) | 0.297**<br>(0.141) | 0.195<br>(0.225) |
| Grades Served (elementary omitted) |  |  |  |
| Middle & high school | -0.354***<br>(0.0799) | -0.461***<br>(0.0825) | -0.688***<br>(0.0829) |
| Not reported | 0.334<br>(0.271) | -0.271*<br>(0.151) |  |
| School Size (<500 students omitted) |  |  |  |
| 500 – 999 students | -0.709***<br>(0.204) | -0.992***<br>(0.142) | -0.611***<br>(0.203) |
| 1000 or more students | 0.269<br>(0.404) | -0.910***<br>(0.245) | -0.814***<br>(0.309) |
| Milk Service Method (in cartons omitted) |  |  |  |
| By dispenser or jug | -0.301**<br>(0.125) | -0.135<br>(0.144) | -1.413***<br>(0.178) |
| Not reported | 0.448***<br>(0.177) | 0.261<br>(0.138) | 0.321<br>(0.138) |
| School in WWF Program | 0.0448<br>(0.135) | 0.0640<br>(0.173) | -0.139<br>(0.200) |
| Interventions Implemented (none omitted) |  |  |  |
| 1 intervention (curricular) | -0.0205<br>(0.107) | 0.0120<br>(0.0937) | 0.0396<br>(0.116) |
| > 1 intervention (curricular + other) | -0.207<br>(0.140) | -0.295***<br>(0.101) | -0.0201<br>(0.108) |
| 1 intervention (non-curricular) | -0.214<br>(0.144) | -0.365***<br>(0.134) | -0.200<br>(0.155) |
| > 1 intervention (non-curricular) | 0.150<br>(0.209) | -0.282*<br>(0.155) | 0.0299<br>(0.244) |
| Time Period (pre-COVID omitted) |  |  |  |
| During COVID (Autum '20 – Spring '21) | -0.463*<br>(0.281) | 0.611<br>(0.405) | 0.121<br>(0.281) |
| Post COVID (Summer '21 or later) | -0.0177<br>(0.199) | 0.418**<br>(0.164) | -0.185<br>(0.163) |
| Season (Spring omitted) |  |  |  |
| Summer | -0.471*<br>(0.262) | -0.337<br>(0.472) | 0.0662<br>(0.368) |
| Autumn | 0.370***<br>(0.123) | -0.0919<br>(0.127) | 0.357**<br>(0.138) |
| Winter | 0.317***<br>(0.0788) | 0.0415<br>(0.0847) | 0.230***<br>(0.0829) |
| Region (Northeast omitted) |  |  |  |
| West | -0.666***<br>(0.144) | 0.406<br>(0.255) | -0.0429<br>(0.204) |
| Midwest | -0.847***<br>(0.157) | 0.0729<br>(0.259) | -0.303<br>(0.210) |
| South | -0.347*<br>(0.194) | 0.879***<br>(0.252) | -0.0121<br>(0.208) |
| Number of entrees served | 0.00892<br>(0.0240) | 0.0365<br>(0.0224) | 0.0318<br>(0.0219) |
| Number of previous audits | 0.00618<br>(0.00624) | -0.00708<br>(0.00795) | 0.00503<br>(0.00666) |
| Constant | -1.965***<br>(0.292) | -3.307***<br>(0.338) | -1.986***<br>(0.296) |
| Observations | 482 | 445 | 427 |
| R-squared | 0.399 | 0.376 | 0.486 |

Notes: Robust standard errors in parentheses. \*\*\* p<0.01, \*\* p<0.05, \* p<0.1.

### School Meal Service Characteristics

Several school meal service characteristics are significantly associated with total per student waste. One core aspect of school meal service involves the length and timing of meals (Figure 3). The regression results suggest that waste is greater when the lunch period begins before 11:00 am (Table 3, column 1). For example, total waste is about 25% lower ([exp(−0.287)-1]*100 = −24.9%, where −0.287 is the regression coefficient) during a lunch period scheduled between noon and 1 pm than during the omitted category (before 11 am, *p* <0.01). This is not significant among older students (Table 3, column 3, *p* > 0.10) nor for produce waste (Table 4, column 2, *p* > 0.10). However, the duration of the lunch period had no significant net association with total waste or any subcomponent, though lunch periods of 30 minutes yield about 17% less total waste among elementary students than when lunch periods are shorter than this (Table 3, column 2, *p*< 0.05). We note that the duration of a school’s lunch period was not reported for the majority of observations, providing us with less confidence in interpreting these results.

**Figure 3.**
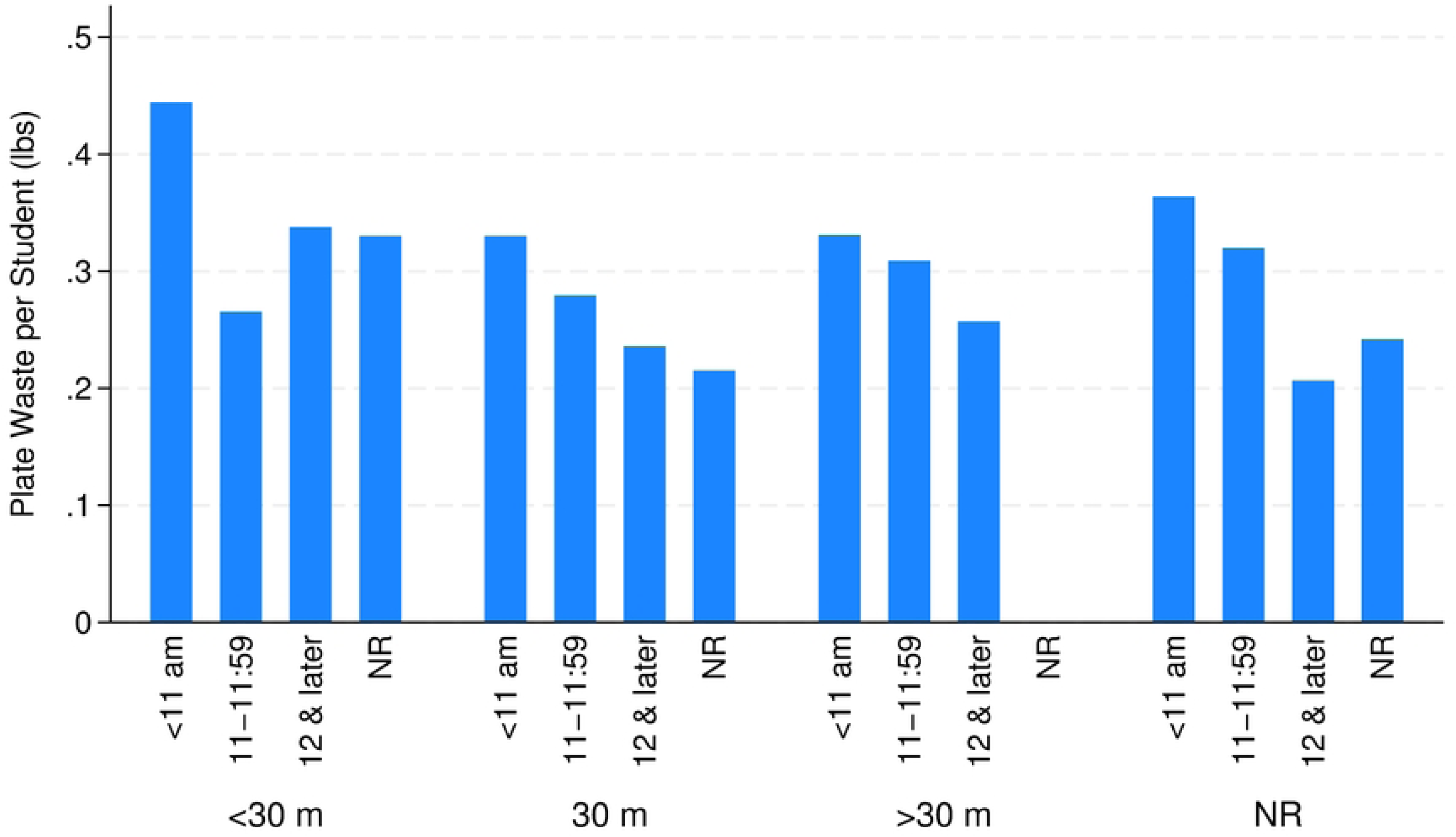
Total Plate and Beverage Waste per Student by Duration and Starting Time of Lunch Period. NR = not reported. The starting time refers to a school’s earliest lunch period for schools with multiple lunch periods. (N=490)

Student autonomy is another aspect of meal service that is postulated to affect waste. We consider two types of autonomy variables observed in this data set: one that provides students with autonomy over which lunch items are placed on their plates and one that permits additional autonomy of the serving size of milk received.

In terms of item-level autonomy, schools may follow two general modes of service: ‘Offer’ in which student may refuse some elements of a standard meal (e.g., take only a fruit or a vegetable), or ‘Serve’ in which all five standard food elements (fruits, vegetables, grains, meat or meat alternative, fluid milk) are provided to every student. Most high schools are required by rule to operate under the ‘offer’ mode of service while it is optional for other grades. In our data, eleven high schools report ‘serve’ mode, which may reflect that the schools are an exception, which can be granted due to insufficient service flexibility, or may reflect schools that have students from both high school and younger grades and the ‘serve’ mode captures service for younger students, or may simply be a miscoding by school volunteers who entered the data.

One would expect that schools implementing an ‘Offer’ mode of service will report less waste as individual autonomy increases permitting students to, e.g., not take a vegetable they are unlikely to consume. However, we find 38% more waste reported in ‘Offer’ schools (coefficient = 0.323, *p*<0.01) with the overall result being driven by middle and high schools (Table 3, columns 1 and 3) with elementary schools having nearly identical waste levels regardless of offer versus serve. These results hold for both categories of food (produce and other solid foods) but not for milk (Table 4).

Shifting to autonomy in the amount of milk served, we find that providing students autonomy to select the amount of milk is associated with 39% reduction in total waste (coefficient = −0.496, *p* < 0.01, Table 3, column 1). While the results are greatest for milk (Table 4, column 3, coefficient = −1.413 or −75.7%, *p* < 0.01) there is also a significant negative association with non-produce solid food waste (26% less).

While each school lunch served must include an entrée to be eligible for federal reimbursement (even if the school adopts the ‘offer’ mode), the school can provide more student autonomy by providing more choices about the entrées selected. Schools that provided more entrée options are marginally significantly associated with more overall waste (∼3% more waste per entrée offered) though the association with each key component listed in Table 4 is insignificant.

### School and Student Characteristics

Young children are associated with greater waste levels than older children with meals at elementary schools yielding about 37% more waste per student than middle or high school students (Table 3, column 1, coefficient = −0.354, *p* < 0.01), where this significant association carries over to each of the three waste components listed in Table 4 with milk waste having the greatest differential across age groups.

Other facets of a school were also observed to significantly correlate to waste per student. School size was one of the largest factors (*p* < 0.001) with schools that served between 500 and 1000 students reporting about 54% less waste than smaller schools, where the association was strongest among schools with older students and carried over to each of the major meal components. The very largest schools (1000+ students served), while not significantly associated with less overall waste, reported significantly less milk and produce waste than the smallest schools.

Schools where approximately half of the school’s population was eligible for free or reduced lunch (25% - 75%) were significantly associated with more total waste than schools with fewer or more students eligible for such benefits. Among meal components the pattern was particularly distinct for produce where schools with the smallest percentage of eligible students had significantly less waste than all other schools (Table 4, column 2). Non-produce waste (e.g., entrees and non-produce sides) were wasted the least in schools with more than 75% of students eligible for free or reduced priced meals.

School region was also significantly correlated with total waste with schools from the Midwest reporting more than 28% less waste per student than those in the Northeast (Table 3 coefficient = −0.321, *p* <0.01). Among older students, the Northeast reported significantly more waste than all other regions (Table 3, column 3). Regional trends were also significant across meal components (Table 4) with entrée waste being significantly greater in the Northeast than all other regions and produce waste being greatest in the South.

The timing of the audit was also significantly associated with waste levels with both timing relative to the onset of the COVID pandemic (*p* = 0.05) and season (*p* < 0.001) implicated. The least waste occurred during the 2020/21 school year (about 51% less than earlier), which coincided with the restart of many schools under restrictive post-COVID meals service routines. In terms of seasons, winter observations yielded about 21% more total waste than spring while entrée and side waste and milk waste was also highest in Fall and Winter. We were also able to document the number of times a school undertook an audit, though the number of previous audits undertaken at that school was not significantly associated with waste level.

The number and type of interventions undertaken at the participating schools revealed some significant association patterns. Compared to the omitted group of schools that undertook no systematic interventions, significantly less waste was observed at schools who undertook a single non-curricular intervention (e.g., focused on behaviors such as sharing uneaten and opened food via share tables, recycling or utensil re-use). However, when considering just produce waste, any intervention that involved at least one non-curricular action was associated with significantly less waste (Table 4, column 2).

## Discussion

Our study yields data collected from 134 schools in 24 states that feature a range of uptake of free and reduced-price meals, different age groups and different meal service models (offer versus serve), and different administrative organization (public, private and charter). Audits were conducted during all seasons and both before and after the onset of COVID. While the data used in this study should not be mistaken for a nationally representative sample, as we rely upon self-reported data from a self-selected set of schools, it does provide a distinct view of plate and beverage waste across a diverse group of U.S. schools. Outside of data collected as part of USDA’s efforts (e.g., School Nutrition Dietary Assessments, see Potamites and Gordon 2010 and Buzby and Guthrie 2002, and the School Nutrition and Meal Cost Study, see USDA 2019), which follow strict sampling protocol and yield nationally representative estimates of plate waste, most other studies of U.S. cafeteria plate waste rely upon observations from cooperating schools collected in a limited geographic region and during a limited time frame (Cohen et al. 2021).

Further, unlike the USDA administered studies, this study relies upon direct measurement of waste amounts via scale rather than indirect measurements that yield the fraction of plated food that remains unconsumed (Buzby and Guthrie 2002, Potamites and Gordon 2010, USDA 2019). While estimates of the proportion of plated food that is wasted are useful, estimates of the physical weight of plate waste can provide useful information for waste management decisions. For example, our estimates suggest that the average elementary student discards 0.236 lbs. of solid food per lunch meal served (Table 4), permitting projections for, e.g., the capacity and throughput requirements of physical infrastructure needed daily should a school consider instituting a composting program. Table 2 also verifies few differences between average per student plate waste amounts created in middle and high schools (see table’s footnote – only milk waste is greater in middle than high schools), though this may be due to limited statistical power, as more comprehensive studies have found differences in the percent of food left on plates between middle and high school students for whole grains, proteins, empty calories as well as milk (USDA 2019, Fig. 5.2).

Several of our results are consistent with suggestions proffered in the literature for schools to reduce plate waste. For example, Buzby and Guthrie (2002) suggest that very early or very late lunch periods may increase waste, and our results confirm that waste was greatest for meals served prior to 11 am with the difference being statistically significant from waste observed during meals served between 12 noon and 1 pm. Also, the waste created during meals between 11 and 12 is significantly greater than those during the noon hour (*p*=0.033).

Buzby and Guthrie (2002) and Cohen et al. (2021) find evidence that very short lunch periods exacerbate plate waste. We find this to hold among elementary students in our sample who are given less than 30 minutes to consume lunch rather than 30 minutes or more (Table 3, column 2), but not to hold for older students (Table 3, column 3). Figure 3 juxtaposes these two factors and suggests that combination of lunch periods that are short in duration (< 30 m) and early in the day (before 11 am) yield the largest reports of per student waste in our sample.

The literature also documents that the timing of lunch periods relative to student recess periods is also a driver of plate waste (Chapman et al. 2017, McLoughlin et al. 2019, Price and Just 2015, Cohen et al. 2021). We note that there was no information provided about the relative order of lunch period and recess, which prohibits us from assessing past research findings of reduced waste during lunch periods that immediately follow periods of physical activity.

We also find results that are partially consistent with the received literature. For example, we find that reported total waste is highest in schools where a moderate fraction of students (25%- 75%) receive free or reduced-price lunches and that milk waste is highest in schools where 75% or more of students receive such benefits. The literature is also mixed on this point with results from USDA’s third School Nutrition Dietary Assessment revealing no significant association for overall waste though significantly more milk waste among marginally food secure students (Potamites and Gordon, pg. 84). USDA’s School Nutrition and Meal Cost Study reveals significantly more waste of both total calories and milk in schools that offered universal free meals versus other schools. Hence, while our results align with the received literature concerning more milk waste in schools with the greatest concentrations of students receiving free and reduced-price meals, we do not align with respect to overall waste. In particular, we find less waste of the main entrée in schools with 75% participation in free and reduced-price meals.

We also find evidence this is distinct from extant studies: schools in our sample that implement the ‘offer’ service mode, in which students need not take all reimbursable meal elements, report more plate waste per student. In contrast, USDA (2019, Table F.15) found lower rates of plate waste for all calories and produce at ‘offer’ elementary schools (but not middle schools) though ‘offer’ middle schools in the USDA study did waste a greater percent of milk than ‘serve’ middle schools. Our results show no significant difference for elementary schools and the opposite pattern of plate waste among schools with older children, i.e., more waste at middle and high schools with ‘offer’ rather than ‘serve.’

There are multiple possible reasons for this discrepancy. First, we rely on absolute levels of waste rather than on the percent of food wasted, opening the door to the possibility that students in offer schools systematically take more total food than those in serve schools and, even though they waste a similar or smaller fraction of food, yield larger absolute waste figures. Second, we rely on a smaller, self-selected sample of schools, where ‘offer’ schools with larger levels of plate waste may have been disproportionately attracted to participate in the study. Third, we are unable to assess offer versus serve status for nearly 12% of the sample, which could alter relative assessments if the unknown schools were correctly assigned to their offer versus serve group. Finally, there is the possibility that patterns have changed in the years since the USDA sample collection ended (2014-15) and our sample collection began (2018-2023). Further research to understand if there is a legitimate driver of more waste in schools using the ‘offer’ service mode is a high priority, as this is a popular option with virtually all high schools and more than 80% of elementary and middle schools operating under this mode (USDA 2019).

Other aspects of meal service that promote student autonomy have mixed associations with waste. Schools in this study that installed dispensers that permit students to select their preferred amount of milk rather than being forced to take an entire carton of milk report 76% less milk waste. While such an outcome is expected and documented in small-scale case studies (WWF 2022), to our knowledge, this is the first documentation in the literature that confirms and calibrates this association. On the other hand, schools that offer more choices of main entrees trend toward more rather than less waste. The literature suggests that providing students more options for individual meal elements is generally associated with less plate waste, as students are more likely to find an option that suits their palate. However, most other studies focus on increasing the number of options available for fruits and vegetables rather than for the main entrée (Cohen et al. 2021).

We also document patterns of waste across schools that, to our knowledge, have not been addressed in previous studies. For example, we find waste is lower in mid-sized schools (500 – 999 students) than in larger or smaller schools, suggesting there may be particular challenges with limiting plate waste for schools operating at the ends of the scale spectrum. Also, we document patterns with respect to seasonality and regionality of plate waste with winter and Northeastern schools associated with significantly greater reported amounts, and with produce waste posing a particular challenge in the South. While the data do not support identifying root causes for such patterns, awareness of these patterns may permit others to identify and address challenges facing schools in different regions and seasons.

Our study is also unique in that schools report any interventions that they have undertaken in their schools to address food waste. Nearly 30% of the observations were taken at schools that had implemented some type of intervention. While we lack the ability to make valid assessments of these interventions, as we do not possess pre- and post-intervention waste levels, we can observe correlations that may be indicative of possible efficacy and warrant further evaluation in more rigorous data settings. For example, we find schools that permit students to portion their own amounts of milk via deployment of a commercial milk dispenser or through self-pouring from jugs report 76% less milk waste than those reliant upon individual milk cartons. Outside of the milk dispenser intervention, schools that implement at least one non-curricular intervention (e.g., a table where students can share unopened, uneaten food items) report significantly less produce waste than schools that either have no interventions or a single curricular intervention. We do not find the act of repeatedly undertaking measurement to be – in and of itself – associated with any more or less plate waste. However, most schools with repeated measurement in our sample tend to have the measurement clustered into a single season, which does not permit much time to respond to any insights the schools may have generated from the act of measurement. Given that measurement is a critical antecedent of action through its ability to diagnose school-level issues and motivate student and staff action, and critical to intervention evaluation, the lack of correlation between repeated measurement and waste levels in this sample should not discourage schools from engaging in measurement.

In general, we lack sufficient detail on the types and intensiveness of the undertaken interventions. When coupled with a lack of data concerning the waste levels before the interventions were deployed, it points to the need for more research to rigorously evaluate the efficacy of different classes of interventions.

## Conclusions

Nearly 7 billion meals reimbursed by the Federal government were served to students in the United States during 2023 (School Nutrition Association 2024). Food from these meals that becomes plate waste represents an opportunity to wield scarce funds more efficiently and support child growth and development more robustly. We analyze data on plate waste from reimbursable lunches recently reported by 134 schools across 24 states to assess commonalities and patterns that might inform efforts to reduce these waste levels. We find the average lunch resulted in 0.34 pounds of uneaten food and beverage per elementary student and 0.23 pounds for other students, suggesting scope for interventions to reduce plate waste, particularly at the elementary level.

Our results are consistent with patterns previously established in the literature, such as greater total waste among younger students and greater milk waste in schools where most students receive free and reduced-price lunches, while inconsistent with some previous work (e.g., we find the most total plate waste among schools with moderate levels of free and reduce-price lunch service rather than the highest levels of such service). We also document patterns not previously explored in the literature, such as higher levels of total waste reported among the smallest and largest schools, schools from the Northeast, and meals served during the winter. While our data is insufficient to assess the root cause of such patterns, this provides guidance for future research that might validate and identify the causal pathways that lead to such patterns.

Our study’s correlative results are also consistent with previously recommended interventions to reduce plate waste, including suggestions to avoid lunch periods that are short and held before standard lunch hours (before 11 am). We also document a significant negative association between providing students autonomy to select their desired amount of milk and the amount of milk that is wasted. While this relationship has been previously hypothesized to hold and documented in small-scale cases studies, we are the first to substantiate this association in a larger sample and calibrate the effect size by finding about a 76% reduction in milk waste.

In some cases, however, we find results that contrast with the received literature, including our finding of that schools with older students operating under the offer versus serve model (permitting students to decline some elements of a reimbursable meal) are associated with more waste despite the additional autonomy this provides to students. As this is an observational study that does not include measurements before and after a school switches modes of service, we caution against drawing causal inferences, but highlight the need to conduct further research to rule out any type of unexpected waste reaction among students in ‘offer’ schools given the prevalence of this mode of service. We also document that other school-initiated waste reduction interventions, particularly those that involve non-curricular activities, are associated with less produce waste. While encouraging, we reiterate the need for more rigorous causal evaluation of such interventions to validate the correlative findings provided by our observational (non-randomized) study.

In addition to the previously mentioned observational nature of the study, we highlight several additional limitations. First, the data set features coverage from only about half of the states in the U.S., and these schools were not randomly selected, but rather represent a group of schools with the motivation and resources to conduct an audit of the waste created in their cafeterias. The data are also lacking information on several key characteristics of the school and meal setting that can inform patterns of cafeteria plate waste, such as the presence of competitive foods (e.g., vending machines or nearby off-campus fast food), whether the school permits students to leave campus for lunch, when recess or other opportunities for physical activity are scheduled relative lunch, and objective measures of the quality of the cafeteria food itself. Future work would benefit from the addition of such variables to the data set.

## Data Availability

All relevant data are within the manuscript and its Supporting Information files.

## Acknowledgements

The authors thank the World Wildlife Fund (WWF) for sharing the data used in this study and Mary Jane Chandler, Alex Nichols-Vinueza, and Pete Pearson of the WWF for helpful conversations concerning the data and initial results and acknowledge the numerous school staff and volunteers who undertook the school food waste audits that constitute the data analyzed in this study. Adjapong recognizes support from the Big 10 Summer Research Opportunities Program and Roe recognizes support from USDA-NIFA (project OHOA-01546) and the National Science Foundation (Grant #2115405). All remaining errors are those of the authors.

## References

Blondin, S., Djang, H., Metayer, N., Anzman-Frasca, S., & Economos, C. (2015). ‘It’s just so much waste.’ A qualitative investigation of food waste in a universal free School Breakfast Program. Public Health Nutrition, 18(9), 1565–1577. Doi:10.1017/S1368980014002948

Burg, X., Metcalfe, J. J., Ellison, B., & Prescott, M. P. (2021). Effects of Longer Seated Lunch Time on Food Consumption and Waste in Elementary and Middle School–age Children: A Randomized Clinical Trial. JAMA network open, 4(6), e2114148–e2114148.

Buzby, J.C. & J.F. Guthrie. (2002). Plate Waste in School Nutrition Programs: Final Report to Congress. Electronic Publications from the Food Assistance & Nutrition Research Program, E-FAN-02, USDA-Economic Research Service, March. Available online at: https://www.ers.usda.gov/publications/pub-details/?pubid=43132 (accessed 17 January 2024).

Byker Shanks, C., Stenberg, M., Bark, K., Izumi, B., Hoff, C., & Parks, C. A. (2023). School Lunch Advisory Councils’ use of behavioural economics influences vegetable selection and waste. Health Education Journal, 00178969231172727.

Capps, O., Ishdorj, A., Murano, P. S., Field, L., Hutto, A., & Storey, M. (2019). Waste Not Want Not. Choices, 34(1), 1–8.

Chapman, L. E., Cohen, J., Canterberry, M., & Carton, T. W. (2017). Factors associated with school lunch consumption: Reverse recess and school “brunch”. Journal of the Academy of Nutrition and Dietetics, 117(9), 1413–1418.

Cohen JFW, Hecht AA, Hager ER, Turner L, Burkholder K, Schwartz MB. Strategies to Improve School Meal Consumption: A Systematic Review. Nutrients. 2021; 13(10):3520. 10.3390/nu13103520.

Hakim, S.M., & Meissen, G. (2013). Increasing Consumption of Fruits and Vegetables in the School Cafeteria: The Influence of Active Choice. Journal of Health Care for the Poor and Underserved 24(2), 145–157. Doi:10.1353/hpu.2013.0109.

Haas, J., Cunningham-Sabo, L., & Auld, G. (2014). Plate waste and attitudes among high school lunch program participants. J Child Nutr Manag, 38(1), n1.

McLoughlin, G. M., Edwards, C. G., Jones, A., Chojnacki, M. R., Baumgartner, N. W., Walk, A. D., … & Khan, N. A. (2019). School lunch timing and children’s physical activity during recess: an exploratory study. Journal of nutrition education and behavior, 51(5), 616–622.

Price, J., & Just, D. R. (2015). Lunch, recess and nutrition: Responding to time incentives in the cafeteria. Preventive Medicine, 71, 27–30.

Potamites, E., & Gordon, A. (2010). Children’s food security and intakes from school meals. Contractor and Cooperator Report No. 2239-2019-2875. United States Department of Agriculture, May. Available online at: https://www.ers.usda.gov/publications/pub-details/?pubid=84339 (accessed 4 August 2023).

School Nutrition Association (2024). School meal statistics. Available online at: https://schoolnutrition.org/about-school-meals/school-meal-statistics/ (accessed on 17 January 2024).

Sharma S, Marshall A, Chow J, Ranjit N, Bounds G, Hearne K, Cramer N, Oceguera A, Farhat A, Markham C. (2019). Impact of a pilot school-based nutrition intervention on fruit and vegetable waste at school lunches. Journal of Nutrition Education and Behavior. Nov 1;51(10):1202–10.

USDA, EPA and University of Arkansas. (undated). “Guide to Conducting Student Food Waste Audits: A Resource for Schools.” Available online at: http://assets.worldwildlife.org/educators_toolkit_files/29/toolkit_files/original/Student_Food_Waste_Audit_FINAL_4-6-2017.pdf?1511297805&_ga=2.261716109.1815262156.1691233805-929868070.1689533680 (accessed 5 August 2023).

USDA (2019). School Nutrition and Meal Cost Study, Final Report Volume 4: Student Participation, Satisfaction, Plate Waste, and Dietary Intakes by Mary Kay Fox, Elizabeth Gearan, Charlotte Cabili, Dallas Dotter, Katherine Niland, Liana Washburn, Nora Paxton, Lauren Olsho, Lindsay LeClair, and Vinh Tran. Food and Nutrition Service, Office of Policy Support, Project Officer: John Endahl. Alexandria, VA: April 2019.

USDA. (2023). National School Lunch Program. Website. Available online at: https://www.ers.usda.gov/topics/food-nutrition-assistance/child-nutrition-programs/national-school-lunch-program/ (accessed 4 August 2023).

Wilkie, A. C., Graunke, R. E., & Cornejo, C. (2015). Food waste auditing at three Florida schools. Sustainability, 7(2), 1370–1387.

WWF (World Wildlife Fund) (2022). “The Business Case for Transitioning to Bulk Milk Dispensers from Single-Use Milk Cartons in K-12 Schools,” December 5. Available online at: https://www.worldwildlife.org/publications/the-business-case-for-transitioning-to-bulk-milk-dispensers-from-single-use-milk-cartons-in-k-12-schools (accessed 27 January 2024).

WWF (World Wildlife Fund) (2023). “Be a Food Waste Warrior” Website. Available online at: https://www.worldwildlife.org/teaching-resources/toolkits/be-a-food-waste-warrior (accessed 5 August 2023).

